# Wakhan: reconstruction of chromosome-scale copy number profiles of tumor genomes with long-read sequencing

**DOI:** 10.64898/2025.12.11.25342098

**Authors:** Tanveer Ahmad, Ayse G. Keskus, Sergey Aganezov, Anton Goretsky, Isabel Rodriguez, Byunggil Yoo, Lisa A. Lansdon, Elena A. Repnikova, Lei Zhang, Yuelin Liu, Ataberk Donmez, Asher Bryant, Sonam Tulsyan, Jimin Park, Joshua Gardner, Brandy McNulty, Samuel Sacco, Jyoti Shetty, Yongmei Zhao, Bao Tran, Salem Malikic, Chi-Ping Day, Karen Miga, Benedict Paten, Cenk Sahinalp, Midhat S. Farooqi, Michael Dean, Mikhail Kolmogorov

## Abstract

A common signature of cancer genomes is a complex, rearranged karyotype, characterized by acquired gains or losses of chromosomal material, referred to as somatic copy number alterations (CNAs). Identification of haplotype-specific CNAs from bulk sequencing data is a key step in many short-read cancer genomic workflows; however, short reads have a limited phasing range. In contrast, long reads can directly phase genomic variants into contiguous haplotypes. Here, we present Wakhan, a long-read method for haplotype-specific CNA calling that can reconstruct longer, up to chromosome-scale CNA profiles of rearranged cancer genomes. Using multi-technology sequencing of a cell line panel, combined with high-quality de novo assemblies, we show that Wakhan CNA profiles have better consistency with sequencing data, as compared to the other popular short- and long-read CNA callers. Further, we show that in combination with accurate somatic SV calls, Wakhan CNA profiles provide additional insights into mutational processes in various breast cancer genomes. Finally, we apply Wakhan to multiple pediatric cancer samples and illustrate the high consistency with standard clinical genetic testing.

## Introduction

A complex and rearranged karyotype is a hallmark of cancer genomes (Cosenza et al. 2022). Various mutational processes result in changes in the number of copies of genomic regions, ranging from small fragments to entire chromosomes, conceptualized as copy number alterations (CNAs) (Krupina et al. 2024). Such alterations could be the result of either focal or chromosome-level losses and gains, as well as more complex processes such as breakage-fusion-bridge (BFB) or extrachromosomal DNA amplifications (Li et al. 2020). As many cancer drivers are explained by CNAs (“Pan-Cancer Analysis of Whole Genomes” 2020), accurate and complete detection of CNA events is a critical step in large-scale cancer genomics studies (Martínez-Jiménez et al. 2023), as well as precision oncology workflows (Sosinsky et al. 2024).

Most current CNA inference algorithms have been developed for bulk whole-genome or whole-exome short-read sequencing technologies (Zhao et al. 2019). While some methods estimate the total number of copies (Babadi et al. 2023), allele-specific or haplotype-specific methods aim to infer CNAs within paternal or maternal haplotypes separately. These methods provide a more precise view of the tumor karyotype and can help to detect additional events, such as copy-neutral loss of heterozygosity (Van Loo et al. 2010).

Commonly used allele- and haplotype-specific CNA methods for short-read sequencing include Battenberg (Nik-Zainal et al. 2012), TITAN (Ha et al. 2014), FALCON (Chen et al. 2015), FACETS (Shen and Seshan 2016), Weaver (Li et al. 2016), CNVkit (Talevich et al. 2016), ReMixT (McPherson et al. 2017), PURPLE (Priestley et al. 2019), HATCHet (Myers et al. 2024) and others. One limitation of these methods is that the phase is typically not preserved between the consecutive CNA calls; thus not achieving chromosome-level reconstruction (Aganezov and Raphael 2020). In addition, SV calling is challenging with short reads (Zook et al. 2020; Wagner et al. 2022), which makes it difficult to associate SVs with CNA boundaries (McPherson et al. 2017). Finally, short-read coverage distribution can be biased due to sampling or mapping artifacts (Benjamini and Speed 2012).

In contrast, long-read sequencing can resolve most common genomic repeats, directly phase germline and somatic variants into long haplotypes, and has better coverage uniformity (Logsdon et al. 2020). Multiple recent studies highlighted the high precision and sensitivity of long reads for somatic SV calling (Shiraishi et al. 2023; Elrick et al. 2025; Keskus et al. 2025), but the application of long reads to haplotype-specific CNA detection has been limited. The algorithms originally designed for short reads can be applied to long-read data (Priestley et al. 2019; Elrick et al. 2025), but they do not take full advantage of long-read phasing or SV calling capabilities.

Here, we present Wakhan, a long-read method that can generate accurate CNA profiles of individual tumor haplotypes. By using long-read phasing information as input, Wakhan can extend the phasing to whole chromosomes by taking advantage of tumor allelic imbalance. Wakhan is also aided by phased somatic SV calls reconstructed by Severus to detect CNA boundaries, resulting in higher precision especially for short CNAs.

We evaluated Wakhan and other methods using a multi-technology panel of CASTLE cell lines (Park et al. 2025) with highly rearranged karyotypes. As currently there are no comprehensive benchmarks for haplotype-specific CNA calls, we designed a CNA evaluation framework enabled by highly-contiguous *de novo* assemblies of matching normal genomes. We show that Wakhan produces CNA profiles that are the most consistent with the sequencing data across technologies. We also illustrate that Wakhan generates better reconstruction of CNA profiles of complex SV rearrangement events, such as chromothripsis, seismic amplification, or breakage-fusion-bridge cycles. We then apply Wakhan to an extended panel of sequenced breast tumor cell lines to discover chromosome-scale rearrangement patterns. Finally, we use Wakhan to characterize rearrangements in several pediatric tumor samples and show high consistency with standard clinical genetic testing.

## Results

### Wakhan algorithm overview

The goal of Wakhan is to generate a haplotype-specific copy number alteration (CNA) profile, where each position of the genome is mapped to two integer copy number states of two parental haplotypes. A complementary goal is to fragment the genome into the maximal contiguous regions with no change in CNA state, which we will refer to as CNA segments. Each non-diploid CNA segment is a result of at least one associated SV breakpoint, with the exception of whole-chromosome gain or loss that does not produce a new breakpoint. To detect CNA segment boundaries, most previous short-read methods primarily relied on detecting the abrupt change in coverage (Olshen et al. 2004), although some methods can use SV calls to refine CNA coordinates (McPherson et al. 2017). Our group previously showed that long-read somatic SV calls are more complete and accurate than short-read SV calls (Keskus et al. 2025). Therefore, Wakhan primarily relies on somatic SV breakpoints as CNA segment boundaries.

Traditional haplotype-specific CNA callers can distinguish haplotypes in regions with an unbalanced number of tumor haplotype copies by analyzing heterozygous variant frequencies, and typically report major and minor tumor CNA states for each CNA segment. However, given two adjacent unbalanced CNA segments, it is unclear if their major alleles are in cis or in trans with respect to true chromosome haplotypes. Wakhan addresses that by considering two key observations: (i) direct long-read phasing often spans CNA boundaries and (ii) a single somatic SV can affect only one tumor haplotype.

Briefly, the algorithm consists of the following parts (Figure 1). Wakhan takes as input a tumor alignment and a vcf with phased germline variants, generated from a matching normal sample alignment, or tumor alignment in tumor-only mode (Supplementary Figure 1). It then splits the genome into bins and computes the median coverage of phased heterozygous SNPs per haplotype. Then, Wakhan synchronizes and extends the input phase blocks in the regions of tumor copy-number imbalance. This phasing extension procedure also generates a new phased vcf file, which is used to produce haplotype-specific SV calls with Severus that define haplotype-specific CNA boundaries. Wakhan then fits the model coverage of one tumor and one normal copy to maximize the fit with the observed coverage. The outputs of Wakhan are a haplotype-specific CNA profile and a small variant vcf with extended phasing. Wakhan also marks CNA segments with substantial deviation from the coverage model as subclonal.

**Figure 1.**
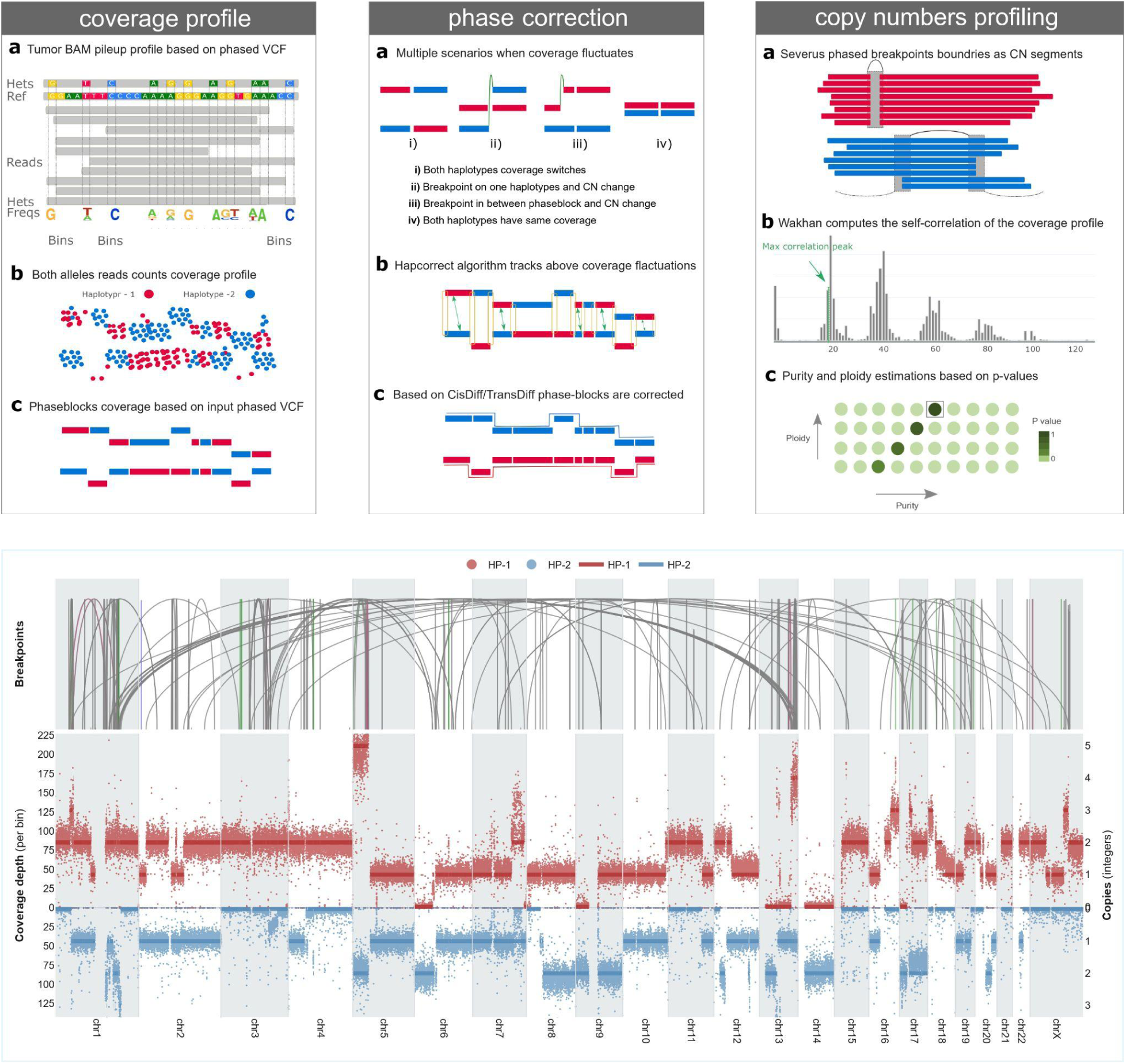
Overview of the Wakhan algorithm and an example of the CNA output. (Top) Key algorithmic steps for the haplotype coverage extraction, phasing correction and CNA profiling. See Supplementary Figure 1 for the complete pipeline. (Bottom) Wakhan CNA output example for the H2009 tumor cell line. Top track shows somatic SVs detected by Severus that are used for CNA boundaries. Bottom track shows inferred CNA profile, as well as observed haplotype-specific coverage. Two haplotypes are shown in red and blue colors.

### An assembly-based framework for CNA profile evaluation

While currently there are several established benchmarks for small variants and SV calling (Zook et al. 2020; Wagner et al. 2022; Fang et al. 2021), we are not aware of any benchmark of similar comprehensiveness for somatic CNA calls. Instead, previous CNA methods have primarily relied on simulations (McPherson et al. 2017), case-by-case analysis (Myers et al. 2024), or inferred ploidy/purity accuracy (Elrick et al. 2025). To enable systematic benchmarking of whole-genome CN profiles with real sequencing data, here we designed a benchmarking framework based on the CASTLE panel, consisting of 6 pairs of tumor/normal cell lines sequenced with multiple short- and long-read sequencing technologies.

While curated sets of haplotype-specific CNA calls are not available for these cell lines, we sought to evaluate CNA accuracy in terms of how well it explains the observed haplotype-specific coverage. Generating reference coverage requires full-chromosome phasing of germline variants, which has been impractical with short-read sequencing. Using long-read technologies, here we generated diploid, highly contiguous de novo assemblies of the matching normal cell lines from the CASTLE panel using PacBio, ONT, Hi-C and PoreC data (Methods). Following the current best practices, we assembled H2009, H1437, and HCC1937 (with PoreC and ultra-long ONT data) using Verkko (Rautiainen et al. 2023), and HCC1954 and HCC1395 (with standard ONT and Hi-C data) using hifiasm (Cheng et al. 2024). This resulted in high-quality assemblies with the median N50 of >130 Mb and median 19+ full-chromosome scaffolds for H2009, H1437, and HCC1937; and N50 = 89 Mb and 9 full-chromosome scaffolds for HCC1954 (Supplementary Table 1). HCC1395 assembly had a reduced contiguity (N50 = 19 Mb), likely due to the lower read N50 of the ONT data; however, the assembly still achieved chromosome-level phasing. Since no poreC or Hi-C data were available for the H578Bst cell line from the CASTLE panel, it was not included in this evaluation.

Given the diploid de novo assemblies, we generated phased SNP variant calls against GRCh38, and computed haplotype coverage of heterozygous SNPs in tumor samples using Illumina, ONT, and PacBio, which we refer to as *reference* coverage (Methods). We define *segment error* (SE) between a CNA profile and a reference coverage as follows. For each identified CNA segment in a chromosome, we compute the mean squared distance between the expected CNA coverage and reference coverage at heterozygous SNP sites for both haplotypes. We then compute the weighted average per chromosome, normalized by the mean coverage of a tumor haplotype (Methods).

The assignment of CNA segment haplotypes into paternal and maternal is initially arbitrary. To compute SE, we match phases of each individual CNA segment and the corresponding reference haplotypes to minimize the error. Thus, SE evaluates the correctness of individual CNA calls, but not the whole chromosome. To measure correctness of CNA profiles on chromosome scale, we introduce a complementary metric - *chromosome error* (CE). It is computed similarly to segment error, but the phase of the entire chromosome - rather than individual CNA segments - is matched against the reference coverage.

### Benchmarking using a multi-technology cell line panel

We selected several state-of-the-art haplotype-specific CNA calling tools to benchmark and compare against Wakhan. For short-read callers, we selected PURPLE (Priestley et al. 2019), Battenberg (Nik-Zainal et al. 2012), and HATCHet (Myers et al. 2024). We also used recently developed SAVANA (Elrick et al. 2025) for long-read CNA calls, and additionally used PURPLE on the long-read data. In the case a tool outputs multiple solutions, we used a solution with the best score. We then used the integer CNA output for each tool. We note that, similarly to the most popular CNA callers (with a few exceptions, such as HATCHet and ReMixT), Wakhan does not explicitly model a mixture of tumor clones, and thus this functionality was not evaluated. For HATCHet, we selected the most abundant clone as representative.

We first computed SE for each tool on the CASTLE dataset against reference coverage produced by all 3 technologies: Illumina, ONT, and PacBio; and we found that the difference in measured error was minimal and highly correlated across technologies (Supplementary Table 3). To avoid circularity in technology comparison, we focused on comparing tools using Illumina and ONT as input against the PacBio reference coverage (all long-read methods support both ONT and PacBio as input). Separately, we compared ONT and PacBio-based inputs against the Illumina reference profiles and confirmed that they were highly similar (Supplementary Table 3).

The median SE was overall reflecting the karyotype complexity of the genomes (Figure 2; Supplementary Figure 2). For example, H1437, with relatively few rearrangements, had median SE ∼0.15 for the best performers, while SE was higher for more complex karyotypes, such as HCC1937 and HCC1954 (∼0.19 and ∼0.23 for best performers, respectively). Note that even for a perfect CNA profile, SE is not expected to reach zero due to fluctuations in coverage depth.

**Figure 2.**
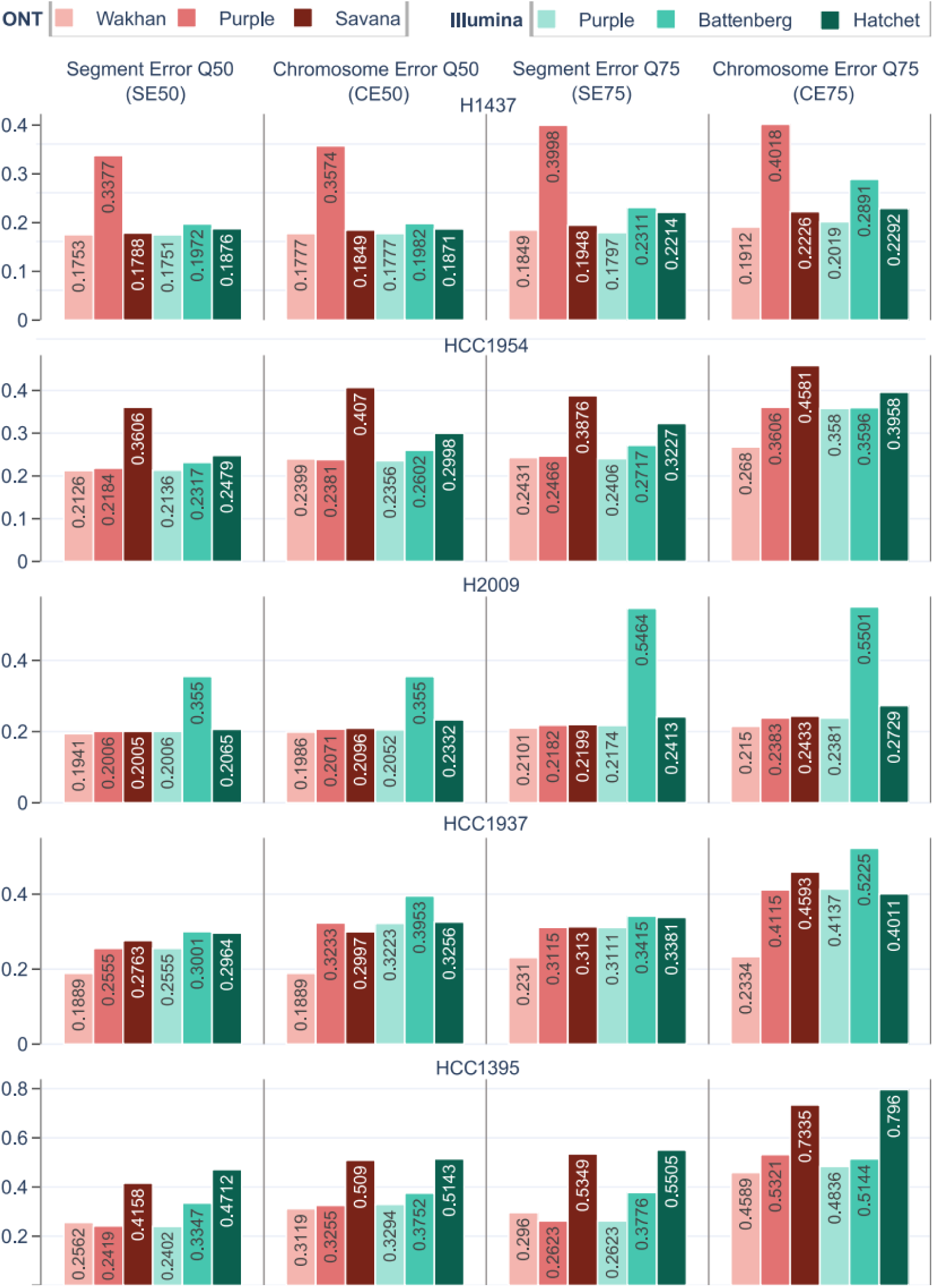
Benchmarking different CNA calling tools using tumor/normal cell lines from the CASTLE collection. Segment error (SE) and chromosome error (CE) are normalized distances between the haplotype CNA profiles output by the tools and the haplotype coverage of PacBio tumor sequencing. SE reflects the error of individual CNA segments, CE measures error of whole chromosome profiles. For both statistics, Q50 and Q75 distributions are shown. Paternal and maternal haplotypes were derived from de novo assemblies of the matching normal genomes.

The median CE was higher than the median SE for all datasets, and the gap was higher for more complex karyotypes. To highlight how different tools handle more challenging chromosomes, we also compared distributions of 75% quantiles of SE and CE. We will refer to the Q50 and Q75 statistics for SE and CE as SE50, SE75, CE50 and CE75, respectively.

Overall, across the five CASTLE datasets, Wakhan and PURPLE showed the lowest SE50 and SE75, compared to other methods, reflecting the high accuracy of individual CNA segment reconstruction (Figure 2; Supplementary Table 2). However, Wakhan consistently had a lower CE50 and especially CE75 metrics, compared to all other methods. This suggests that Wakhan produced a more accurate representation of chromosome-scale CNA profiles, compared to all traditional haplotype-specific CNA tools, especially for highly rearranged chromosomes.

### Wakhan generates an improved representation of chromosome-scale CNA profiles

The improvements in Wakhan CNA profiles, compared to other tools, were the most pronounced in chromosomes with complex CNA profiles and rearrangement architecture (Figure 3). In our analysis, we observed two main patterns that explain the improvement. First, Wakhan produced a better reconstruction of chromosomes with many clustered SVs, typically resulting from complex mutational processes, such as chromothripsis, seismic amplification, or breakage-fusion-bridge. These processes are expected to affect a single chromosome and thus should be phased into a single haplotype. For example, in HCC1954, Wakhan reconstructed a complex seismic amplification of the chr8q arm, likely targeting MYC, coinciding with loss of the p-arm on the same haplotype (Figure 3a). While Wakhan attributed most CNA changes to a single haplotype, other tools split the CNA between two haplotypes.

**Figure 3.**
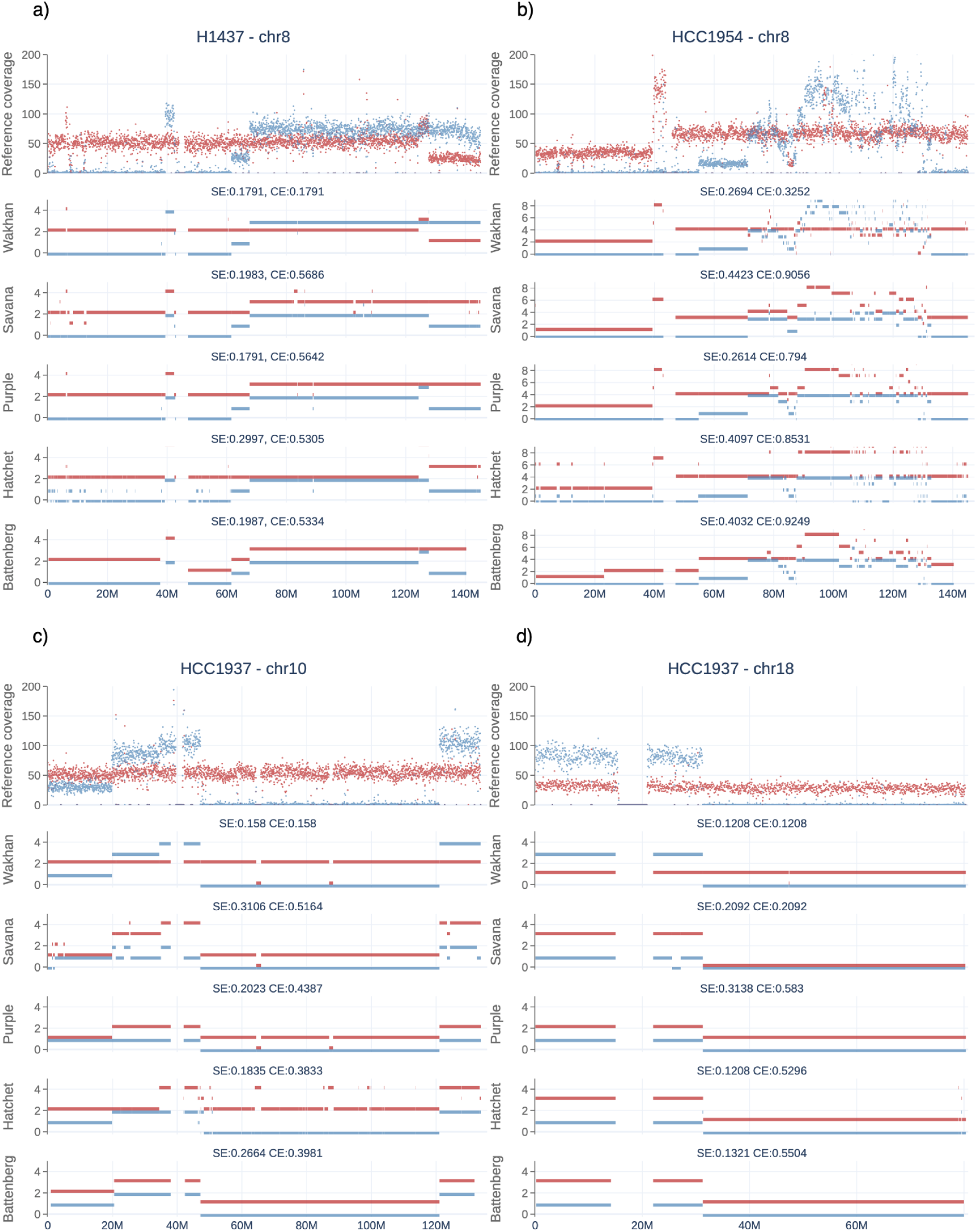
Examples of chromosome-scale reconstruction of complex karyotypes by Wakhan and other methods. The top of each panel shows reference coverage for two haplotypes (in red and in blue). CNA profile outputs of each tool are given below. Each CNA profile is annotated with segment error (SE) and chromosome error (CE).

The second challenging scenario where Wakhan outperformed other tools was chromosomes with large amplifications and losses affecting the same haplotype (Figure 3b-d). In contrast to Wakhan, other tools often attributed amplifications and losses to different haplotypes.

Overall, we observed a substantial number of examples where Wakhan reconstructed more accurate chromosome-scale profiles, compared to other tools. We defined challenging cases as chromosomes for which CE was at least 20% higher than SE. In highly rearranged genomes (HCC1954, HCC1937, and HCC1395), we detected 6, 8 and 6 challenging cases in other tools that were correctly resolved by Wakhan. More stable genomes (H1437 and H2009) had 2 challenging cases each (complete CNA profiles visualization for all CASTLE datasets available in Supplementary Data). Overall, this highlights that Wakhan can provide accurate reconstructions of highly rearranged chromosomes that are challenging to resolve without phasing.

### Benchmarking with variable tumor purity and sequencing depth

While the cell lines above have challenging and rearranged genomes, an orthogonal source of complexity in real tumor samples analysis is reduced tumor cell purity (normal contamination) and lower sequencing depth. To test the robustness and accuracy of different tools in these conditions, we generated synthetic mixes of matching tumor and normal cell lines with variable tumor cellular purity and total coverage. We simulated purity for 60% and 80% DNA content, which translated to ∼30% and ∼50% cellular purity because of higher cancer ploidy levels. The normal sample was downsampled to 30x depth. We selected HCC1954 and H1437 genomes for downsampling experiments, representing relatively high and low-complexity rearrangements, respectively. In cases where Wakhan outputs two solutions with very similar scores (within 1%), we selected the one with ploidy matching the majority of all outputs.

For both Wakhan and PURPLE Illumina, SE and CE overall remained stable despite the reduction of tumor purity and sequencing depth, showing robustness in increasingly challenging scenarios (Figure 4; Supplementary Tables 4-7). In HCC1954, Wakhan CE was slightly increasing with the decrease in coverage and purity; however it remained the lowest compared to all other tools. In H1437, the changes in Wakhan and PURPLE chromosome error were minimal. Other tools were less stable with an increased number of synthetic datasets with reduced quality, as compared to 100% purity with maximum coverage. We also compared the inferred purity and ploidy numbers, which overall showed the expected consistency and were less informative, compared to SE and CE statistics (Supplementary Figures 3-4).

**Figure 4.**
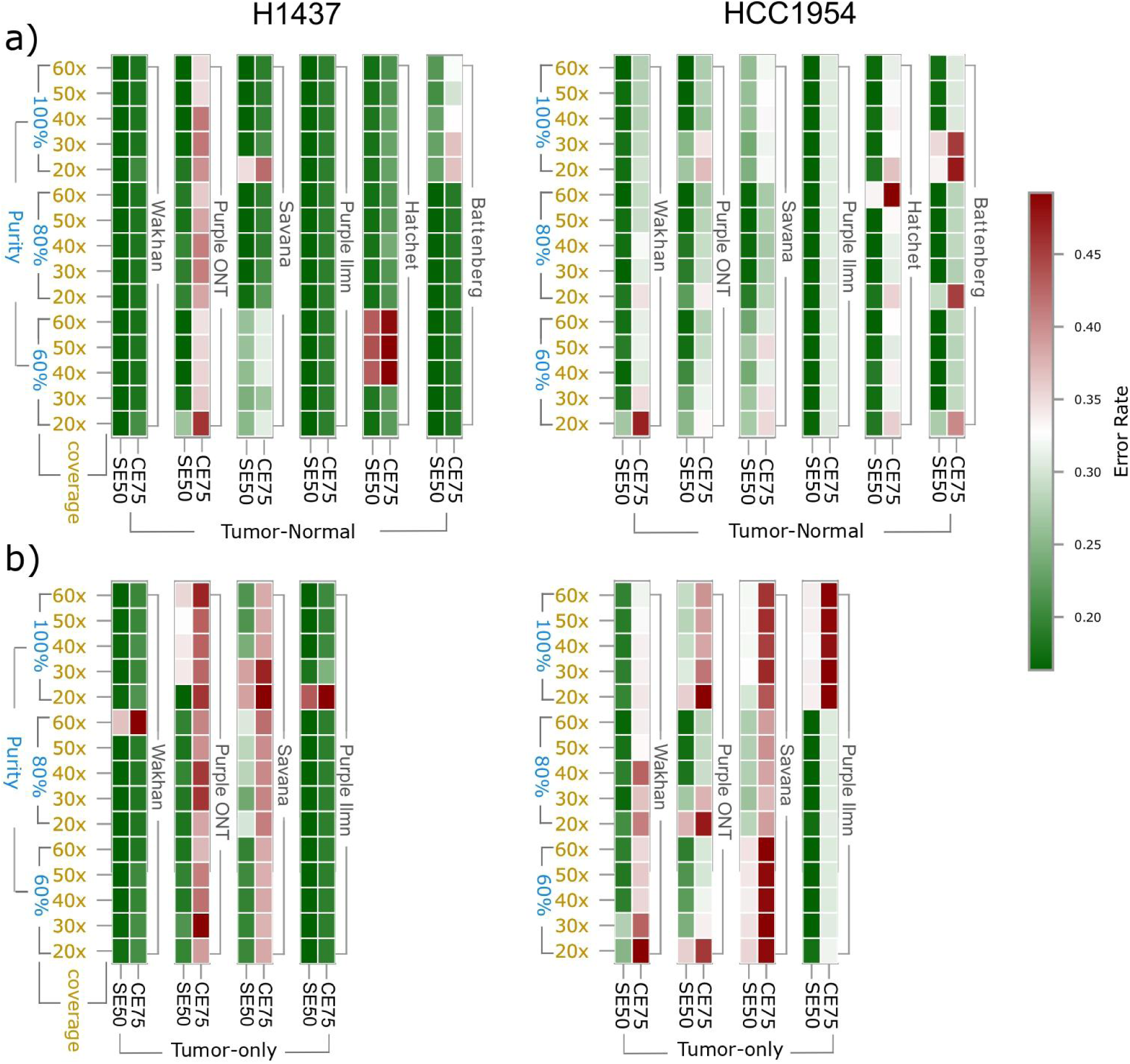
CNA profile error rates for synthetic HCC1954 and H1437 mixtures with variable purity and sequencing depth. Each method was run on a grid of synthetically mixed tumor/normal samples. Tumor DNA fraction ranged from 60x to 100x, and total sample coverage ranged from 20x to 60x. Segment error Q50 (SE50) and chromosome error Q75 (CE75) are shown. **(a)** Tumor-normal runs are shown at the top, and **(b)** tumor-only runs on the bottom.

### Benchmarking tumor-only CNA profile reconstruction

Matching normal samples is not always available, and thus, we tested Wakhan, PURPLE, and SAVANA in tumor-only mode. HATCHet and Battenberg currently do not provide support for tumor-only runs and thus were not tested in this section. Similarly to the analysis above, we used the synthetic mixtures of HCC1954 and H1437, but only with the tumor mixture as input.

Overall, both Wakhan and PURPLE were robust to the absence of normal samples and showed good correspondence with the tumor/normal profiles (Figure 4; Supplementary Tables 4-7). On HCC1954, Wakhan had slightly increased SE and CE, due to several chromosome regions designated as loss of heterozygosity (LOH); however, CE remained lower, as compared to all other methods. On H1437, all tools produced results similar to those of tumor/normal runs.

### CNA profiles of 8 breast cancer genomes provide insights into amplification mechanisms

We next analyzed patterns of chromosomal rearrangements in an expanded panel of breast cancer genomes. In addition to 3 cancer cell lines in the CASTLE panel (HCC1954, HCC1937, and HCC1395), we sequenced 5 more breast cancer cell lines: BT474, MDA-MB-361, MDA-MB-453, SKBR3, and ZR7530 (Methods). The extra cell lines were sequenced using a single ONT flow cell, with a median coverage of 27x and median read N50 ∼30kb (Supplementary Table 9). Matching normal samples were not available for these cell lines, and thus analysis was run in tumor-only mode.

Wakhan reconstruction of haplotype CNA profiles revealed both recurrent and distinct patterns of chromosome-scale amplifications, losses, and rearrangements (Figure 5). For example, we observed recurrent LOH on chromosomes 1p, 3p, 8p, 9p, 13q, and 17p, which have been commonly described in breast cancer (“Comprehensive Molecular Portraits of Human Breast Tumours” 2012). Out of 86 observed cases, ∼60% affected parts of chromosome arms, while whole arms were deleted in ∼33% cases. Finally, in ∼7% cases LOH affected entire non-acrocentric chromosomes, deleting both arms; interestingly, half of all full-chromosome LOH cases were detected in HCC1937. In this cell line, all but 3 chromosomes had at least partial LOH, and many retained haplotypes were present in 3-4 copies.

**Figure 5.**
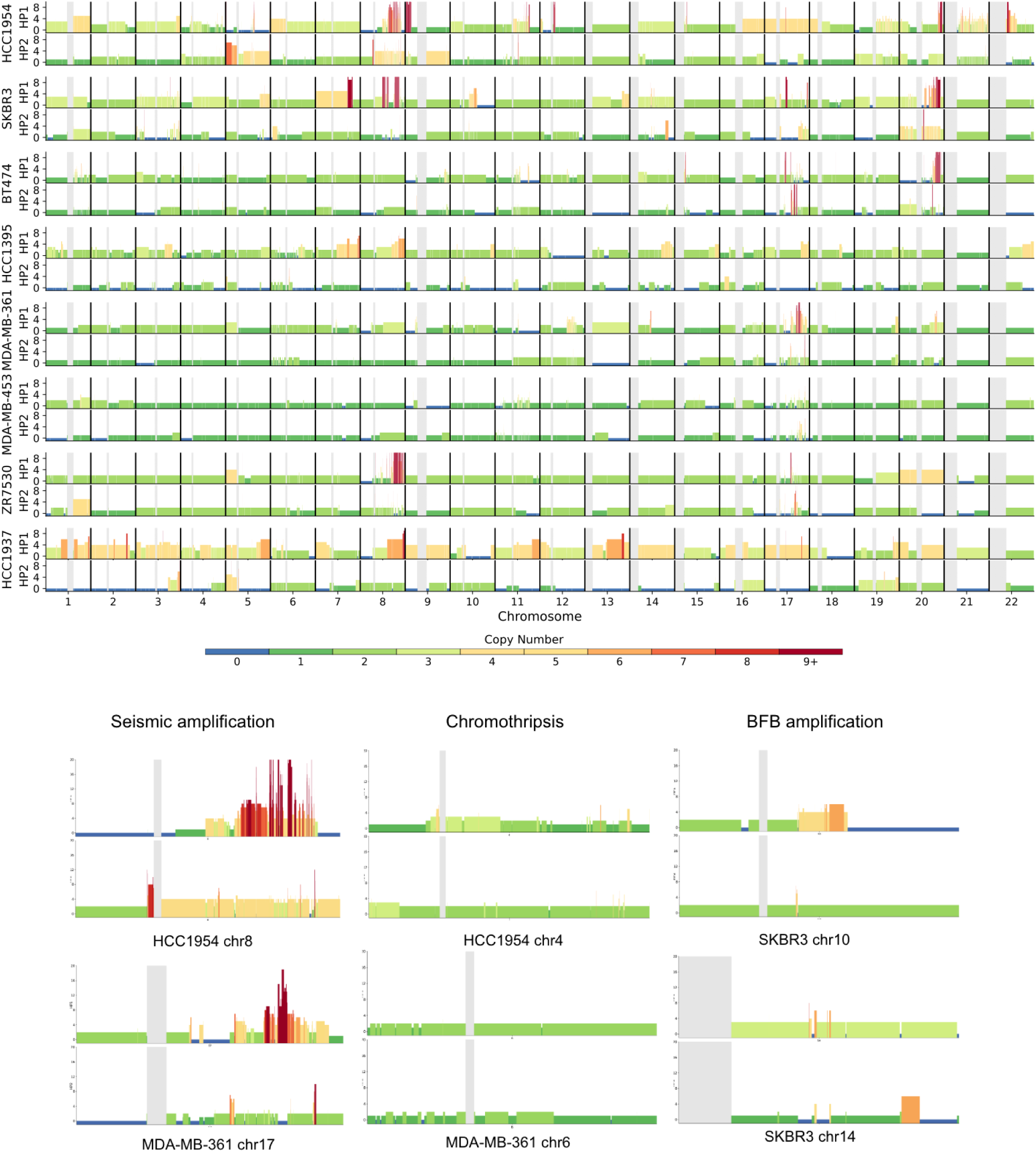
Chromosome CNA profiles for 8 breast cancer cell lines reconstructed by Wakhan. (Top) Visualization of Wakhan haplotype-specific CNA profiles for each breast cancer cell line. Haplotype 1 is assigned as the haplotype with the highest mean coverage for each chromosome. Copy numbers are shown with bar height and color legend. (Bottom) Examples of CNA signatures of different mutational processes. Haplotype-specific amplification profiles of relevant genes from the COSMIC database (Sondka et al. 2024) are shown on Supplementary Figure 6.

We next looked at the patterns of apparent complex amplifications of chromosome haplotypes, characterized by a higher proportion of CNA segmentation and an increased number of copies per segment (Supplementary Table 10). As expected, in most cases such patterns affected only one chromosome haplotype, suggesting that the corresponding rearrangements were acquired in a single burst (Stephens et al. 2011). We observed recurrent high copy number (>20) amplifications in chromosomes 8q, 17q, and 20q, which are among the most common amplifications in breast cancer. CNA profiles of these chromosomes were consistent with seismic amplification patterns, characterized by a high number of segments, fluctuating CNA profiles, and a large number of copies (Rosswog et al. 2021). These patterns were distinct from chromothripsis CNA profiles, recurrently observed in chromosomes 4, 6, 9, and 13; characterized by longer segments and fewer CN states (“Criteria for Inference of Chromothripsis in Cancer Genomes” 2013). This was further different from BFB amplifications observed in chromosomes 6 and 10, characterized with stair-like amplifications, followed by a telomeric LOH in the same haplotype (Zakov et al. 2013).

Overall, this analysis shows that haplotype-specific CNA profiles (Figure 5), complemented by high-quality SV calls, are well-suited for studying the mechanisms of genome rearrangements.

### Wakhan CNA profiles of pediatric tumor samples show high consistency with clinical panels

Since the analysis above was performed using tumor cell lines as input, we next aimed to validate the performance of Wakhan on long-read sequencing of blood and solid tumor samples. Three pediatric tumor samples were selected from Children’s Mercy Hospital: acute myeloid leukemia (CMH-AML), diffuse midline glioma (CMH-DMG), and osteosarcoma (CMH-OS). These patients underwent standard genomic testing as a part of their clinical care, which included chromosome analysis, genomic microarrays, FISH analysis, and short-read whole-exome sequencing for small variant discovery. For this study, we sequenced three retroactive samples to represent karyotypes with low, medium, and high degree of complexity, based on the available data from genomic panels. Tumor and matching normal samples for DMG and OS, as well as a tumor-only sample of AML were sequenced using PacBio with median coverage 47 and median read N50 ∼11 kb (Supplementary Table 11).

For the CMH-AML sample, Wakhan output CNA profiles suggesting a near-diploid karyotype, with gain of chromosomes 8 and 21 (Figure 6). Additionally, terminal losses of 7p and 7q were observed, which Severus predicted to form a ring chromosome. These findings were consistent with the clinical chromosome, FISH, and microarray findings. The chromosome analysis and microarray also demonstrated gain of chr19; however, Wakhan only observed a segmental gain around the centromere.

**Figure 6.**
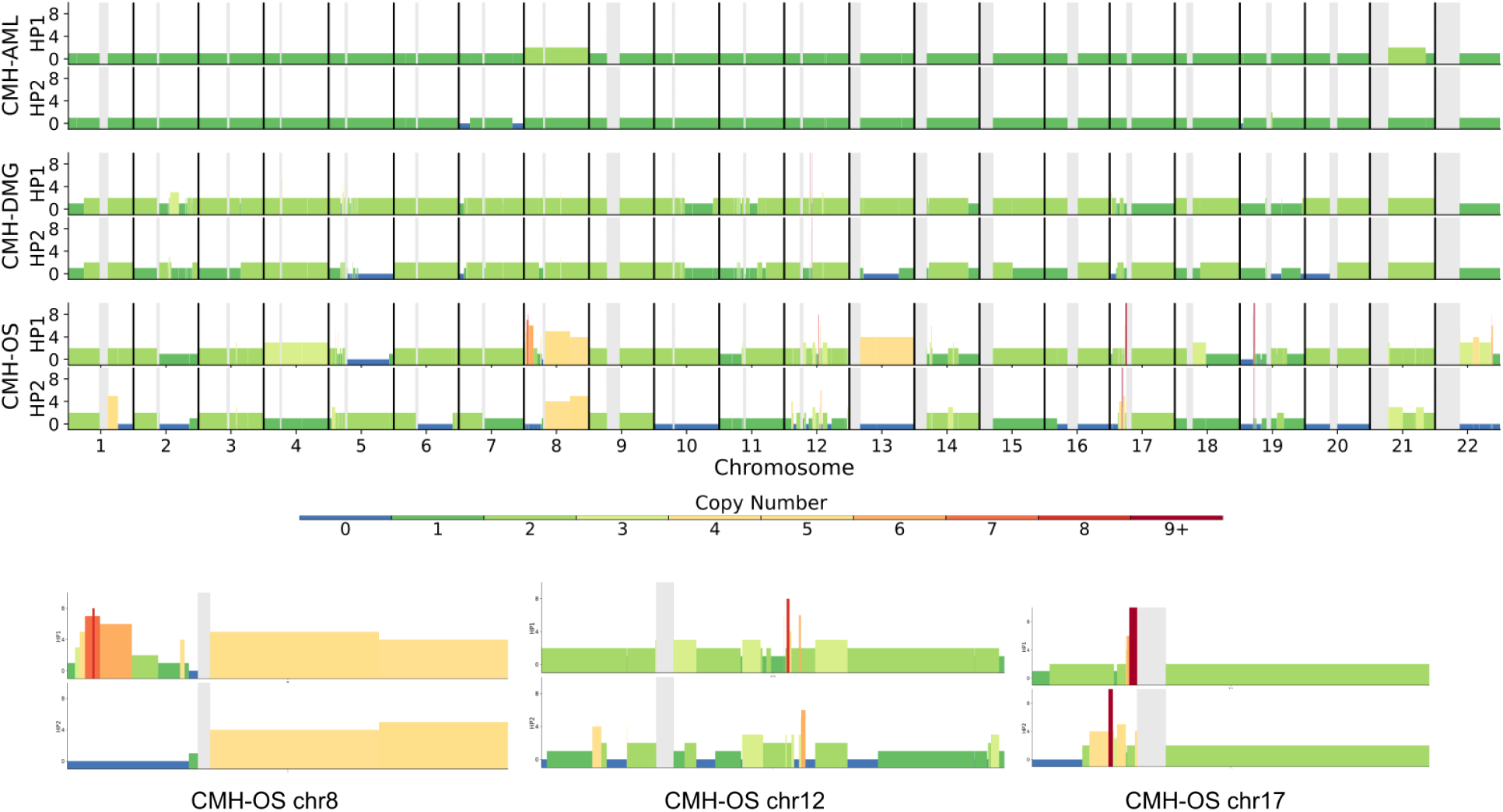
Wakhan CNA profiles of clinical pediatric cancer samples with various degrees of karyotype complexity. (Top) Profiles were reconstructed from clinical PacBio sequencing of pediatric acute myeloid leukemia (CMH-AML), diffuse midline glioma (CMH-DMG), and osteosarcoma (CMH-OS). (Bottom) Examples of 3 most rearranged chromosomes from the CMH-OS sample.

For the CMH-DMG sample, Wakhan reported a more complex karyotype consisting of several structurally abnormal chromosomes with segmental gains varying from three to four copies and multiple regions of LOH (Figure 6). These findings were highly consistent with the clinical cytogenetic results which demonstrated a complex near-triploid karyotype with microarray detecting that most chromosomes were present in 3 to 4 copies with additional segmental gains and losses. The microarray also detected losses involving chromosomes 3p, 5q, 7p, 11p, 11q, 13q, 17p, and 20p, as well as chromothripsis of chromosomes 5 and 11, all of which were largely consistent with the Wakhan and Severus results.

Finally, for the CMH-OS sample, Wakhan output an even more complex karyotype, with approximately half of chromosomes affected by rearrangements or LOH, with chromosome 13 in copy-neutral LOH (CN-LOH) state (Figure 6). These findings were supported by the clinical cytogenetic results which demonstrated a complex hypo-tetraploid karyotype with multiple rearrangements and losses in roughly half of the chromosomes. The microarray additionally detected CN-LOH of chromosome 4, and demonstrated considerable chromosomal complexity including loss of chromosomes X, 10, 13, 16, 20, 22 and the majority of 5q, gain of chromosome 14, and segmental gains and losses of chromosomes 1, 2, 3, 6, 8, 12, 19 and 21, highly consistent with Wakhan CNA profiles. In addition, Wakhan identified additional complexity within chromosomes 5p, 8p, 12, 17p, 19, 21, 22, as compared with the prior microarray testing. The presence of foldback inversions and stair-like rearrangement patterns within chromosomes 8, 17 and others suggested that they were affected by BFB amplifications, while others - such as 12 - had the signature of chromothripsis (Figure 6).

Overall, the analysis of these three pediatric cancer samples of varying cytogenetic complexity showed that Wakhan CNA profiles were highly consistent with prior clinical genetic testing and could potentially be used as a complementary diagnostic tool in personalized oncogenomics.

## Discussion

In this work, we presented Wakhan, a tool for generating haplotype-specific CNA profiles from long-read tumor sequencing data. In contrast to currently available short- and long-read tools, Wakhan leverages long-read connectivity to preserve haplotype phase beyond CNA boundaries and reconstruct more contiguous, often chromosome-scale CNA profiles.

We compared Wakhan against the other popular short- and long-read CNA inference methods using a panel of five tumor/normal cell lines. To enable the benchmarking, we generated diploid *de novo* assemblies of matching normal cell lines and used germline variants from these assemblies to calculate the reference coverage of tumor haplotypes. We then demonstrated that Wakhan generated more accurate chromosome-scale CNA profiles compared to all other tools, particularly in highly rearranged genomes. We also illustrated that Wakhan is robust against varying tumor purities and sequencing depths, and performed well in tumor-only mode.

We then demonstrated the benefits of the phased CNA approach on a panel of eight breast cancer cell lines. Wakhan reconstructed CNA profiles of various degrees of complexity, consistent with the known recurrent amplification sites in breast cancer, but providing additional insights into distinct amplification and rearrangement mechanisms. Accurate CNA and SV analysis enabled by long reads is critical for accurately characterizing complex rearrangement karyotypes commonly observed in cancer genomes, and their role in cancer evolution.

We further demonstrated the application of Wakhan to real tumor samples by generating CNA profiles of three pediatric tumors with varying degrees of karyotype complexity. Wakhan CNA calls were highly consistent with prior clinical genomic testing, suggesting that personalized oncology workflows could benefit from long-read analysis integration. Our groups are currently validating Wakhan and Severus for the genetic diagnosis of prospective samples in an expanded patient cohort from Children’s Mercy Hospital (Lansdon et al. 2024).

Similar to most benchmarked CNA methods, Wakhan does not explicitly model clonal mixtures; instead, it flags subclonal regions that deviate from the coverage model. This works well for dominant clones but may miss signals in polyclonal tumors. However, unambiguous clonal deconvolution requires multiple matching tumor samples as input (Myers et al. 2024), which is not typically produced in clinical workflows.

Wakhan requires phased germline SNP calls as input with phasing block length of hundreds of kb or longer. This is typically achieved in either PacBio or ONT sequencing of tumor biopsies (Lansdon et al. 2024; O’Neill et al. 2024). While the phasing extension algorithm mitigates fragmentation and errors, limited phasing quality due to low read length and shallow sequencing depth can present a challenge. Tumor-only phasing presents an additional difficulty due to high allelic imbalance and tumor clonality; therefore, new algorithms designed specifically for phasing tumor genomes are likely to improve phasing quality (Ming-En Ho, Zhenxian Zheng, Ruibang Luo, Huai-Hsiang Chiang, Yao-Ting Huang 2025).

We note that some short-read methods can use phasing with population databases to enhance the signal from individual SNPs (Hofmeister et al. 2023). However, the linkage information quality quickly degrades over the genomic distance and thus becomes unreliable for chromosome-scale analysis. In principle, reference panel phasing can be complemented by long-read phasing to enhance the phasing length. We also note that single-cell genomics methods, such as CHISEL (Zaccaria and Raphael 2020) and Alleloscope (Wu et al. 2021) have many conceptual similarities to the methods benchmarked in this study; however evaluation of single-cell methods is beyond the scope of this study.

Our CNA benchmarking framework is among the first attempts to design a systematic benchmark for CNA profiling tools using real sequencing data. It is, however, limited to cell lines, which may not represent the whole complexity of the real heterogeneous tumors. Real tumor benchmarks, on the other hand, face the issue of sample availability and data access. High consistency between Wakhan CNA profiles and prior clinical genomic testing for three patient tumor samples provides an encouraging start and expanded validation cohort studies are currently ongoing (Lansdon et al. 2024).

## Acknowledgements

This research was supported in part by the Intramural Research Program of the National Institutes of Health (NIH). The contributions of the NIH authors are considered Works of the United States Government. The findings and conclusions presented in this paper are those of the authors and do not necessarily reflect the views of the NIH or the U.S. Department of Health and Human Services. This work utilized the computational resources of the NIH HPC Biowulf cluster (https://hpc.nih.gov).

## Methods

### Ethics statement

For the three pediatric tumor samples, participants were enrolled by Children’s Mercy Hospital into its institutional Tumor Bank research study, which was approved by the Children’s Mercy Hospital Institutional Review Board and included participant consent for the collection, processing, storage and sequencing of samples. For cell line sequencing, the Institutional Review Board of the National Institutes of Health considers patient-derived cell lines as nonhuman subjects; thus, approval was not required.

### Wakhan overview

In the following sections, we describe the key parts of the Wakhan method. As input, Wakhan takes tumor and normal long-read alignments in BAM format, and a set of phased germline SNP calls in a vcf format. In our benchmarks, we used Clair3 (Zheng et al. 2022) to generate germline SNP calls and LongPhase (Lin et al. 2022) for phasing; however, Wakhan can work with any other suitable methods. Wakhan first synchronizes and extends the input phasing by taking advantage of haplotype coverage imbalance. This produces extended, often chromosome-scale phasing if there is sufficient allelic imbalance. Then, Severus is run with the extended phasing vcf as input to produce phased SV calls. These calls are then used as input for the Wakhan CNA module that fits a haplotype coverage model assigning integer copy-number states to CNA regions (A complete pipeline illustration is given in Supplementary Figure 1).

The main Wakhan outputs are haplotype-specific CNA profile and vcf file with extended phasing. Wakhan produces several additional outputs with expanded information about CNA regions and interactive visualizations. If there are several alternative CNA solutions with close likelihood scores (for example, with or without whole-genome duplications), Wakhan outputs multiple alternative solutions, and the user can select one that is more biologically plausible.

### Generating haplotype coverage and identifying switch errors

Below we will denote two germline haplotypes as haplotypes A and B (since the true parental state is unknown). Given phased heterozygous SNPs and a tumor alignment, Wakhan first computes haplotype-specific coverage in each SNP position, and then aggregates the coverage into 10-50kb bins. In addition, Wakhan computes the median coverage of haplotypes A and B within each phased block. While long-read phasing is typically accurate, it may produce occasional switch errors. If a phase block covers the region of haplotype imbalance, Wakhan detects points where binned haplotype coverage inverses, and breaks such phase blocks at the inversion coordinate.

### Haplotype phasing extension algorithm

Current haplotype-specific CNA inference algorithms implicitly phase CNA segments by assigning major haplotypes (with higher coverage) in each block to haplotype A, and minor haplotype as B. While this works in some cases, as we illustrated in the Results sections, min/maj haplotyping strategy often results in erroneous CNA reconstruction in more complex and rearranged chromosomes.

Direct short- and long-read phasing methods can phase heterozygous variants into contiguous phasing blocks. Recent studies have shown that long-read phasing often achieves megabase-scale phasing block lengths (Lin et al. 2022; Shafin et al. 2021); however, to achieve chromosome-scale phasing, additional data, such as Hi-C, poreC, or pedigree information, is required (Cheng et al. 2022). In contrast to balanced diploid genomes for which most phasing tools were designed, cancer genomes often have an imbalance in the number of paternal and maternal chromosome copies, which could be leveraged to extend initial phasing blocks. Intuitively, this could be achieved by synchronizing the haplotypes of adjacent phasing blocks to minimize the number of CNA state changes in each haplotype.

The algorithm input is a set of phasing blocks P_1_, P_2_ .. P_n_, each with two coverage values Cov_a_(P_n_) and Cov_b_(P_n_), corresponding to the median coverage of the haplotypes *A* and *B* in phase block P_n_. We also denote *LCov_a_(P)* and *RCov_a_(P)* as flanking left and right coverage of the block’s haplotype A. Given two adjacent phase blocks P_1_ and P_2_, the goal of a *block flipping* operation is to decide if the respective haplotypes A and B in these blocks are in *cis* or in *trans* (with respect to the true germline haplotypes).

The Wakhan block flipping algorithm works as follows. Given two adjacent phase blocks P_1_ and P_2_, we consider three major cases: (i) there is no somatic CNA between the beginning of P_1_ and the end of P_2,_ (ii) there is one or multiple CNA boundaries inside either P_1_ or P_2_, or both, and (iii) there is a CNA boundary in between P_1_ and P_2_, Note that if we assume that germline phasing boundaries and somatic CNA boundaries are independent, case (iii) should be relatively rare.

In the case (i), since there are no CNAs in the region between the start of P_1_ and the end of P_2_, true coverage of haplotypes A and B remains unchanged; therefore, we should flip blocks so that Cov_a_(P_1_) = Cov_a_(P_2_) and Cov_b_(P_1_) = Cov_b_(P_2_). Case (ii) is reduced to case (i) if we consider the flanking coverage of P_1_ after the rightmost CNA boundary and the flanking coverage of P_2_ until the leftmost CNA boundary. In practice, if the CNA boundary is close to the phase block boundary, measuring the flanking coverage might be difficult, and the case is processed as case (iii) instead. In case (iii), we assume that one somatic CNA changes the coverage of one haplotype; therefore, the coverage of another haplotype remains unchanged. Thus, to decide if blocks P_1_ and P_2_ are in cis or trans, we compute:

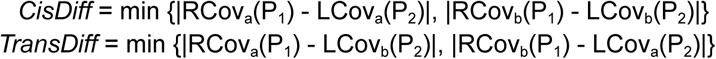

If *TransDiff* is lower than *CisDiff*, P_2_ is flipped with respect to P_1_. Note that this formula extends to the cases (i) and (ii) as well, and therefore we use the same formula for all the cases.

In practice, phase blocks with uncorrected erroneous phasing and short phase blocks with uncertain coverage may present difficulties for the flipping procedure. To account for that, after the first round of flipping Wakhan finds and merges contiguous phase blocks that have consistent haplotype imbalance between adjacent pairs. Short singleton blocks that were not merged this way are removed, and then a second iteration of block flipping is performed on merged blocks. In the second iteration, we use a variation of the formula above to put additional weight on the CNA-changing haplotype:

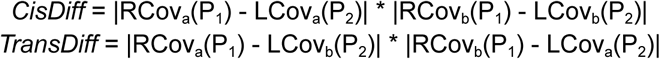

Through empirical search, we found that 5Mb typically works well as a flanking distance for phase block coverage. It typically corresponds to entire phase blocks on the first iteration, but benefits the second iteration with much longer merged phase blocks.

To validate accuracy of extended phasing, we compared N50, switch and hamming error rates of phasing before and after Wakhan extension (Supplementary Table 8; Supplementary Figure 5). In this comparison, assembly-based phasing was used as ground truth. The comparison showed that both switch and hamming errors remained similar or lower, while phasing N50 increased by an order of magnitude.

### Integer copy-number inference algorithm

The haplotype correction procedure above generates a new vcf file, which is used by Severus to produce haplotype-specific SV calls. These SVs serve as CNA boundaries in the corresponding haplotype. If an SV is unphased (is outside of any phase blocks), this SV breaks both haplotypes. We also add annotated centromere start and end as additional CNA boundaries, as SVs in centromeres are still challenging to detect.

Using binned haplotype coverage, Wakhan generates a haplotype coverage distribution (Figure 1). In this distribution, each peak corresponds to a different number of tumor copies with a normal cell admixture, e.g. Cov = xT + N, where T and N are the coverage of one T and N haplotype, respectively, and x is the number of tumor copies (could be zero for a region with a loss of heterozygosity). The next step of the algorithm aims to fit T and N from the observed distribution.

To fit T, Wakhan computes the self-correlation of the coverage profile (correlation against itself) with a range of linear shifts, and T is selected as a shift with the best correlation value. Note that the N value does not affect this calculation since the correlation coefficient is invariant with respect to linear transformations. After inferring T, Wakhan is optimizing N via a grid search from 0 to T. For every putative N, Wakhan scores the corresponding coverage model as the consistency between the observed and modeled coverage per segment. The local maxima reached during the grid search are then output as solution candidates.

We highlight that while Wakhan optimizes T and N coverages, the other CNA methods typically optimize purity and ploidy via grid search. Since ploidy and purity could be computed given T and N, the corresponding search problems are in principle equivalent, although in practice noise may be affecting the two search strategies differently. Another difference is that Wakhan performs two sequential optimizations of T and N, while purity/ploidy inference methods typically perform a full search in a 2D parameter space.

### Tumor-only mode

In the case that matching normal samples are not available, Wakhan can run in the tumor-only mode. We first perform variant calling and pairwise phasing of the tumor sample, which brings additional challenges. First, somatic variants create additional tumor haplotypes; however, since germline variants are much more frequent compared to somatic variants, pairwise phasing algorithms collapse tumor haplotypes together with corresponding germline haplotypes. Secondly, a large imbalance in haplotype coverage can confuse phasing algorithms if it expects a balanced read support of two haplotypes. In practice, we found that LongPhase was robust to most of these challenges in the regions outside of extreme allelic imbalance and coverage fluctuations.

Regions of LOH in the tumor may remain unphased if the normal coverage in this region is low (or zero in the case of 100% pure tumor cell lines). Therefore, all variants in these regions will be unphased. In tumor-only mode, Wakhan detects these regions with very low heterozygous SNP density. Then, since LOH regions only contain one tumor allele, Wakhan treats all SNP variants as heterozygous with the phase assigned to a single arbitrary haplotype, which is then synchronized with the rest of the chromosome during the phase correction stage.

### De novo assemblies of normal CASTLE cell lines

To generate de novo assemblies of matching normal samples BL2009, BL1437, and HCC1937BL, we used PacBio, ONT, and PoreC data as an input for Verkko 2.2.1 (Rautiainen et al. 2023), as it is currently the only assembler that natively supports PoreC data. The resulting assemblies had very high contiguity with NG50 over 130 mb, and on average 19 full-chromosome scaffolds (Supplementary Table 1). Since poreC data were not available for HCC1954BL and BL1395, we downloaded publicly available Hi-C data (Brunette et al. 2024; Fang et al. 2021). We tested both Verkko and hifiasm 0.25.0 (Cheng et al. 2024) to assemble both genomes, and selected hifiasm assemblies for the further analysis since they were slightly more contiguous. HCC1954BL was a highly contiguous assembly (NG50 = 91 Mb) of comparable quality to the poreC-phased assemblies. For BL1395, contiguity was lower (NG50 = 21 Mb), which is likely due to substantially shorter ONT read length of this data (N50 = 11kb), as compared to other normal cell lines (read N50 = 40-60 kb). However, even with reduced contig length, Hi-C data provided chromosome-scale phasing. Both assemblers were run using default parameters.

We performed QC of the resulting assemblies with QUAST, yak, and asmgene, which confirmed the high quality of the resulting assemblies, comparable to the state-of-the-art assemblies produced in other studies (Supplementary Table 1).

We then aligned the diploid assemblies against GRCh38 and used dipcall to generate phased small variant calls. Afterwards, we computed haplotype-specific coverage of each heterozygous SNP in the matching tumor sequencing data using “samtools mpileup”. We finally removed variants inside annotated centromere regions. This SNP coverage data (produced with either ONT, PacBio, or Illumina) was then used to benchmark CNA calls.

We also visualized and manually verified the resulting coverage profiles per chromosome to ensure the quality of phasing. Large-scale switch errors are visually apparent from the coverage profile in the regions of haplotype imbalance, corresponding to abrupt inversions in coverage of two haplotypes. We observed on average two potential large-scale phase switches per genome for BL2009, BL1437, HCC1937BL, and HCC1954. BL1395 assembly unexpectedly had eight apparent phase switches, which may be a consequence of reduced assembly contiguity. However, it is also possible that phase switches are explained by early homologous recombination in tumor genomes and need to be further explored. We note that these apparent disagreements between the tumor and normal phasing affected all benchmarked tools similarly.

In addition to putative switch errors, we found large regions with LOH in two chromosomes in each of BL1437, BL2009, and BL1395. The corresponding tumor genomes did not harbor these losses, suggesting that they occurred exclusively in normal cell lines after they were established.

### CNA benchmarking framework

We benchmarked the output of CNA profiles of various tools as follows. The input of a benchmark is (i) a CNA profile in bed format, consisting of chromosome segments with integer tumor copy number pair *(CopyA, CopyB)* for each segment *Seg,* referred to as *modelled* copy numbers, and (ii) haplotype-specific coverage of heterozygous germline SNPs by tumor sequencing, denoted as *reference* coverage.

First, we fit a simple linear model to establish the correspondence between the CNA copy-number state and expected coverage ModCov(CopyA) ∼ CopyA to maximize the fit with the reference coverage profile. The model has a null intercept (since there is no normal coverage in the reference data), and its slope coefficient corresponds to the coverage of a single tumor haplotype in the reference data. We note that the vast majority of CNA profiles converged to the same slope coefficient within each dataset (Supplementary Table 2).

To compute *segment error (SE)*, for each CNA segment with *(CopyA, CopyB)* and corresponding haplotype reference coverage (Cov_a_, Cov_b_) we first synchronize the phase of the CNA and reference haplotypes to maximize the similarity. We then compute SE for CNA haplotype as the mean square difference between *Cov_a_* and *ModCov(CopyA)* for each SNP position. SE for chromosome is a weighted average of CNA segment errors, normalized by the mean reference haplotype coverage.

*Chromosome error (CE)* is computed using a similar procedure, but instead of synchronizing the phase of individual CNA segments, we synchronize the phase of the entire chromosome.

### Downsampled synthetic datasets generation

Synthetic mixes of matching tumor and normal of both cell lines, HCC1954 and H1437 for multi-technologies (Illumina, ONT and PacBio) were generated for different purity titration and coverage downsampling. As mentioned above, the normal sample was downsampled to 30x depth for all technologies. We produced 100%, 80% and 60% DNA tumor purity mixtures and then downsampled each mixture to 60x, 50x, 40x, 30x, and 20x coverage depth. We used samtools to downsample tumor BAMs with fixed *seed=5* for downsampling pure tumor normal pair samples, while *seed=55* was fixed for downsampling the merged (mixed) BAM files with given tumor fraction. The full script is available in the supplementary archive (see Data availability).

### Running other CNA tools

Wakhan (version 0.3.0), Purple (version 4.1), Savana (version 1.3.2), Battenberg (version 2.2.9) and HATCHet (version 2.1.1) were run in tumor and normal pairs mode using default settings. Similarly, for tumor-only mode we run Wakhan, Purple and Savana with the same versions. For Wakhan, we used “*Phased SVs/Breakpoints pipeline mode” by* enabling phased structural variations from the Severus option using *--use-sv-haplotypes.* Briefly, to use phased breakpoints Wakhan works in two steps, in first step it uses phase-switch correction module to rephase VCF, which is used to haplotag BAMs through WhatsHap v2.8 (Martin et al. 2016), then Severus uses haplotagged BAMs to generate phased breakpoints. In the second step, Wakhan takes this resultant Severus phased SVs VCF and runs the copy number module. Wakhan and Savana require phased germline VCF in tumor and normal pair mode while Wakhan also requires phased germline tumor VCF in tumor-only mode, so we used Clair3 (version v1.0.5) (Zheng et al. 2022)) to generate germline SNP calls and LongPhase (version v1.6) ((Lin et al. 2022)) for SNPs phasing with default settings for clinical samples, similarly for CASTLE cell lines and breast cancer cell lines we used PEPPER-Margin r0.8 (Shafin et al. 2021) for variant calling and WhatsHap for phasing as mentioned in Severus. Hatchet was run by setting *phase_snps=FALSE* to get more contiguous copy number segments. In Purple, we set min/max purity and ploidy to 0.1/1 and 1/8 respectively. A detailed description of commands/scripts is given in the supplementary archive (see Data availability). Complete workflow is available via Lumos pipeline (https://github.com/KolmogorovLab/Lumos).

### ONT sequencing of five breast cancer cell lines

The cell lines (BT474, MDA-MB-361, MDA-MB-453, SKBR3, and ZR7530) were obtained from the American Type Culture Collection (ATCC). Cells were cultured in EMEM or RPMI-640 media with 10% fetal bovine serum, 1% Pen-Strep (10,000 units/mL of penicillin, 10,000 μg/mL of streptomycin, and 25 μg/mL of amphotericin B) until 70%–80% confluent. Cells were washed with 10 mL PBS and harvested using 2 mL trypsin per T-75 flask. All cell lines were confirmed by Identifier analysis and were regularly tested for mycoplasma infection. DNA was extracted and purified using a Gentra Puregene kit from Qiagen. DNA was quantified by Qubit (Thermo Scientific) and stored at 4°C.

DNA was size-selected using the PippinHT system (Sage Science, USA). DNA with >75ng/ul and up to 1500ng/lane was processed, and four lanes per cell line were run to yield enough DNA for two LSK114 libraries. The manufacturer protocol was followed using a size cutoff of 15kb on HPE7510 cassettes. The DNA obtained from the PippinHT was quantified using qubit and TapeStation.

The Ligation Sequencing Kit (SQK-LSK114, Oxford Nanopore Technologies (ONT)) was used for long-read DNA sequencing of the cell line DNA samples using a previously described protocol (Malik et al. 2023). To target an N50 of about 30kb and about 20-30x coverage, the DNA was size-selected. Libraries were loaded on PromethION R10.4.1 flow cells on a P2 sequencer. Flow cells were washed and reloaded to reach about 20-30x coverage per cell line using the ONT Wash kit (EXP-WSH004).

### PacBio sequencing of pediatric cancer samples

Sample libraries were prepared and sequenced as previously described (Lansdon et al. 2024). Libraries were sequenced on the Sequel IIe or Revio System on one or two SMRT cells for a targeted depth of 30-60x coverage.

## Code availability

Wakhan is freely available at https://github.com/KolmogorovLab/Wakhan under the MIT license.

## Data availability

CASTLE data is available under project NCBI SRA BioProject PRJNA1086849, and alternatively from https://github.com/CASTLE-Panel/castle. Additional ONT sequencing is being uploaded to SRA, and clinical sequencing data will be available through dbGaP. De novo assemblies, CNA reference profiles, benchmarking scripts and outputs of all benchmarked tools are available at: https://doi.org/10.5281/zenodo.17780981.

## Supplementary Figures

**Supplementary Figure 1.**
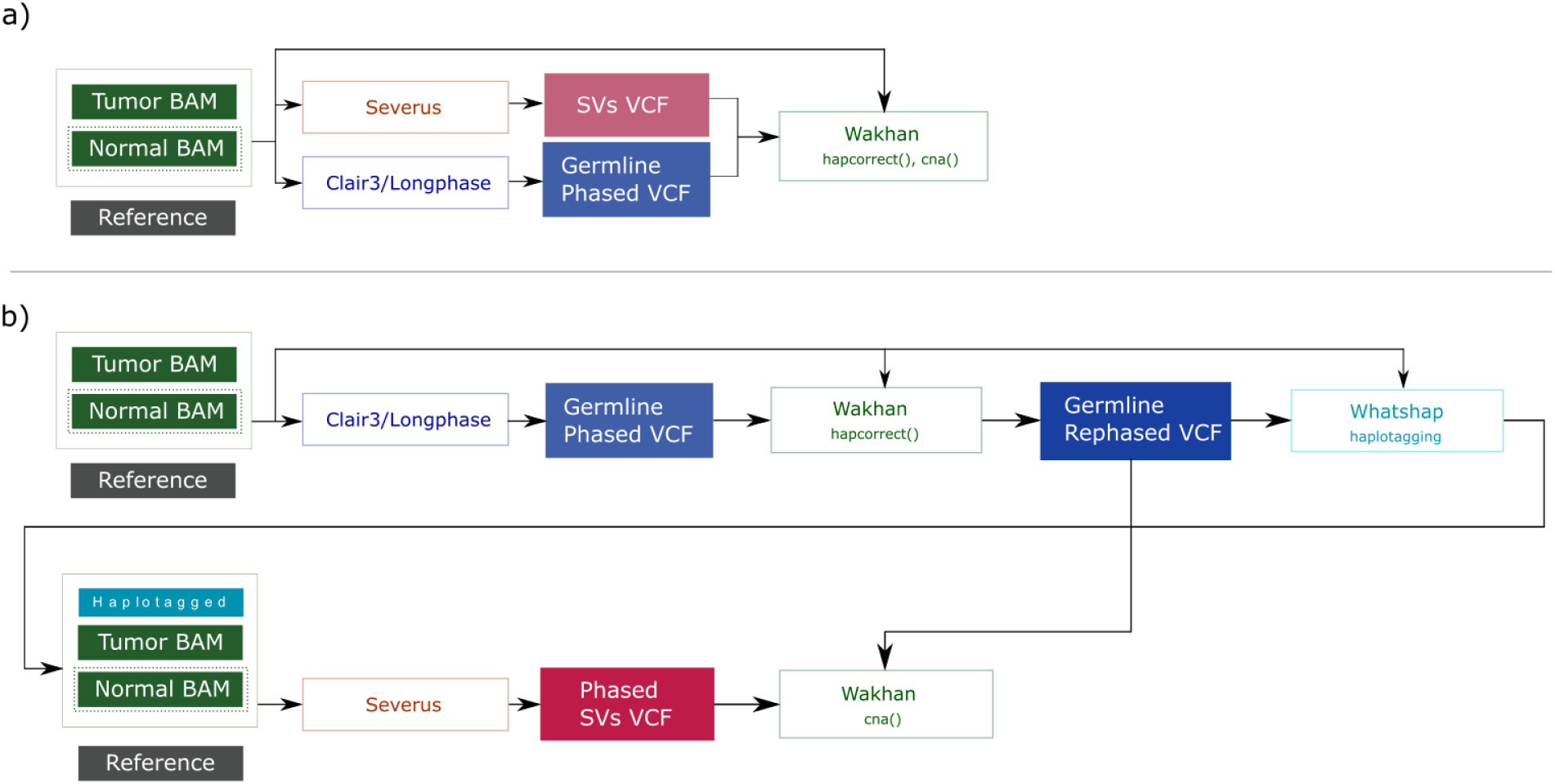
Full Wakhan pipeline. Wakhan can be run as a **(a)** standalone phase correction and copy number profiling tool. In case phased SVs/breakpoints are available from Severus, it could be run as **(b)** phased SVs/breakpoints pipeline mode.

**Supplementary Figure 2.**
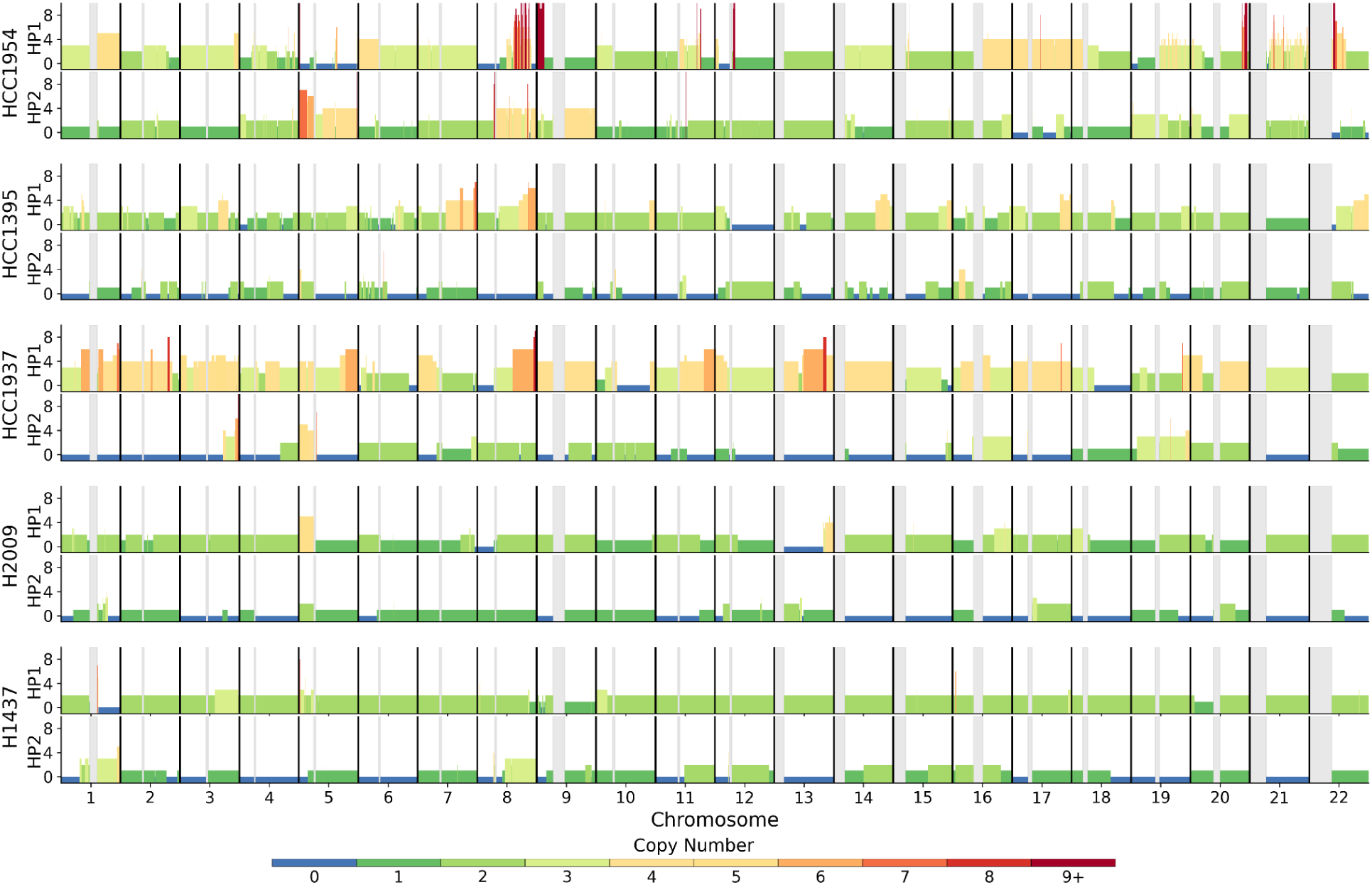
Wakhan CNA profiles of tumor genomes from the CASTLE panel.

**Supplementary Figure 3.**
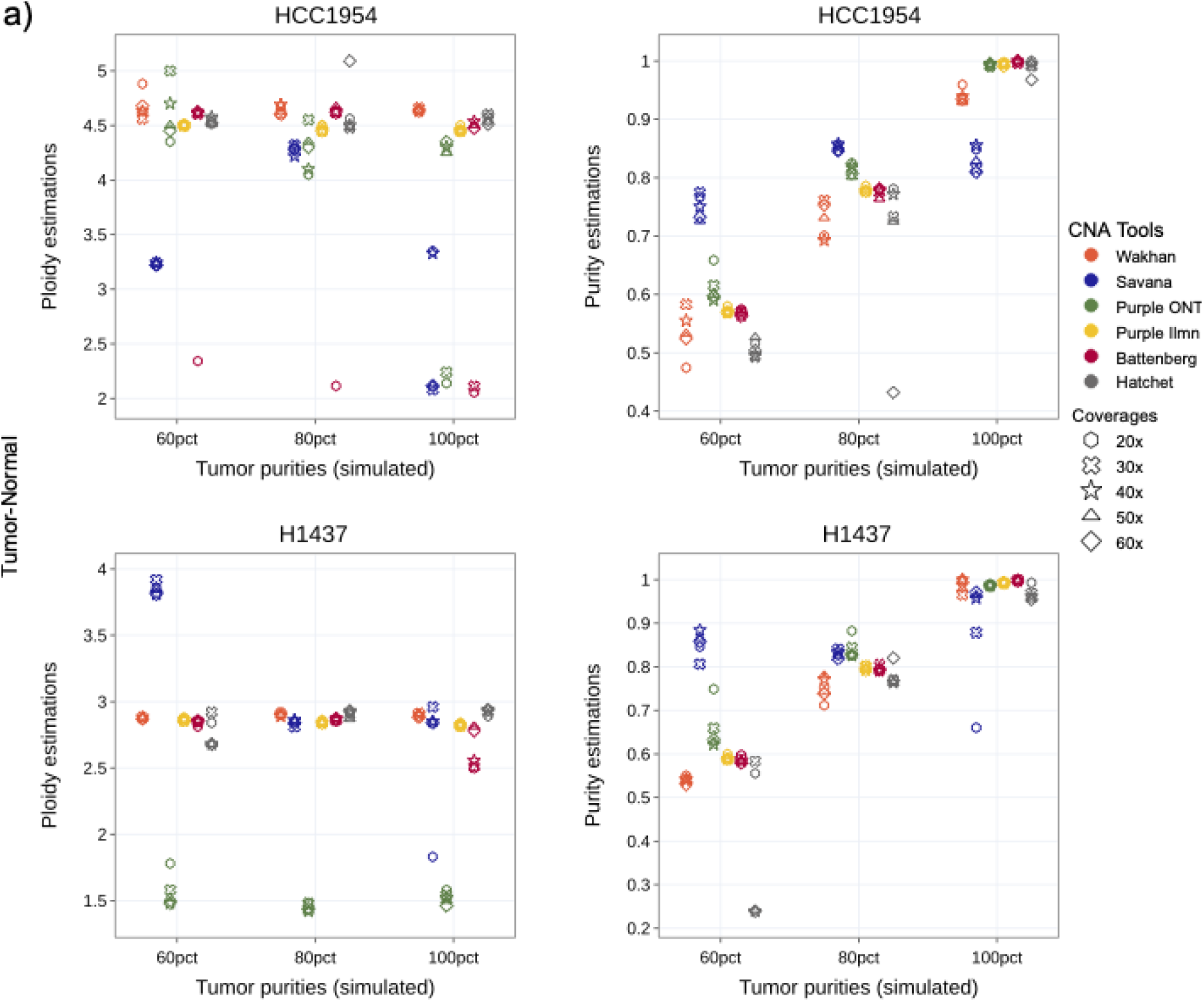
Ploidy and purity inference accuracy on downsampled datasets in tumor-normal mode.

**Supplementary Figure 4.**
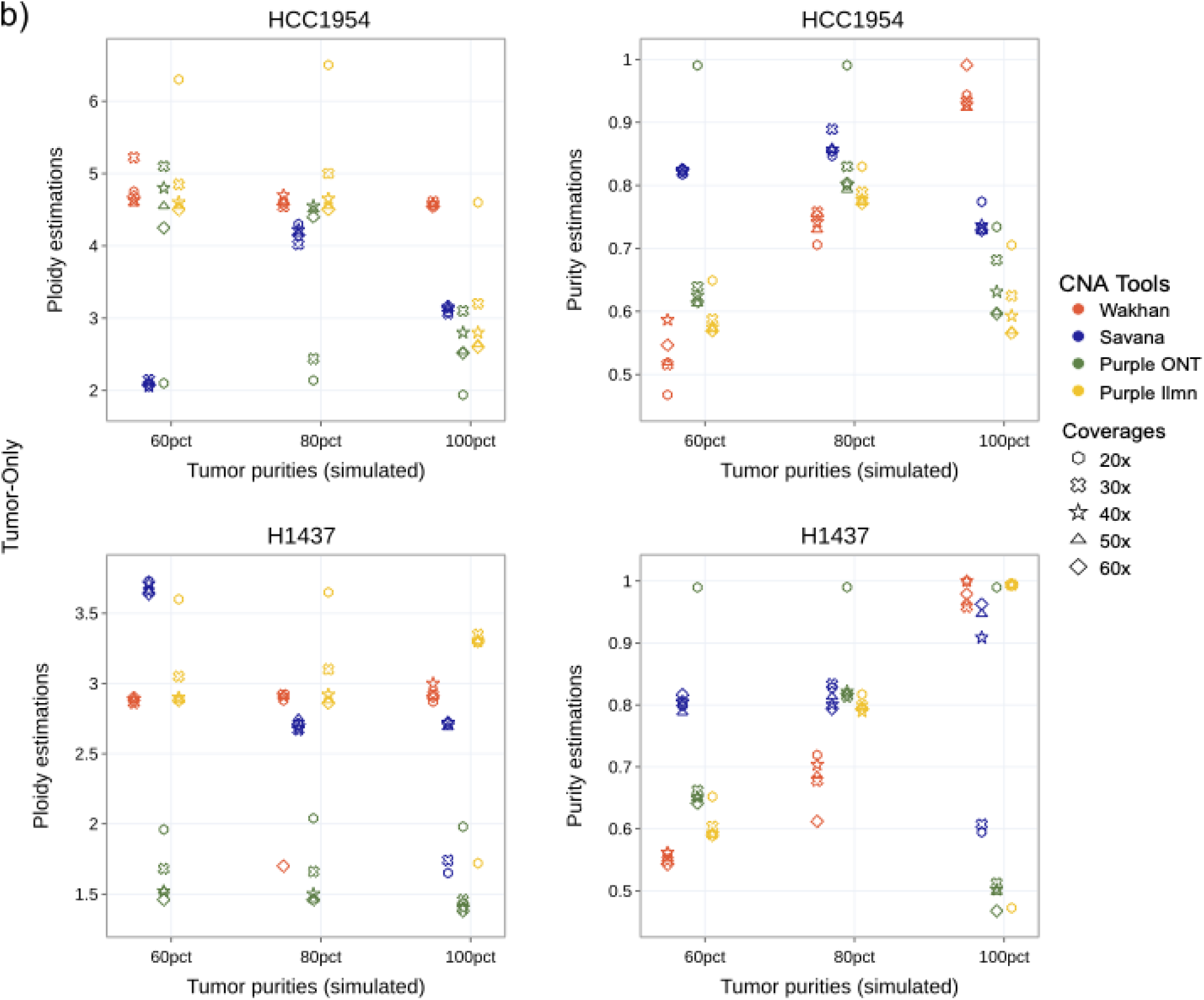
Ploidy and purity inference accuracy on downsampled datasets in tumor-only mode.

**Supplementary Figure 5.**
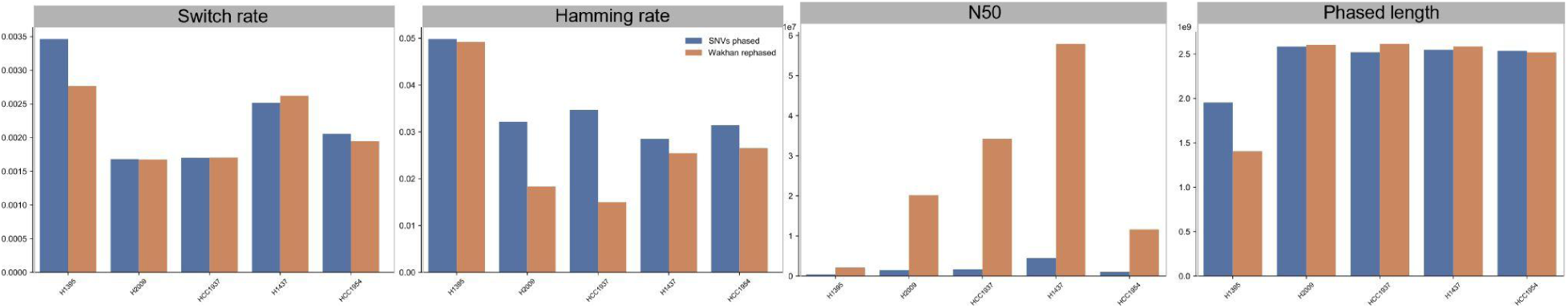
Phasing statistics (Switch rate, Hamming rate, N50 and phased length) before and after Wakhan phasing correction.

**Supplementary Figure 6.**
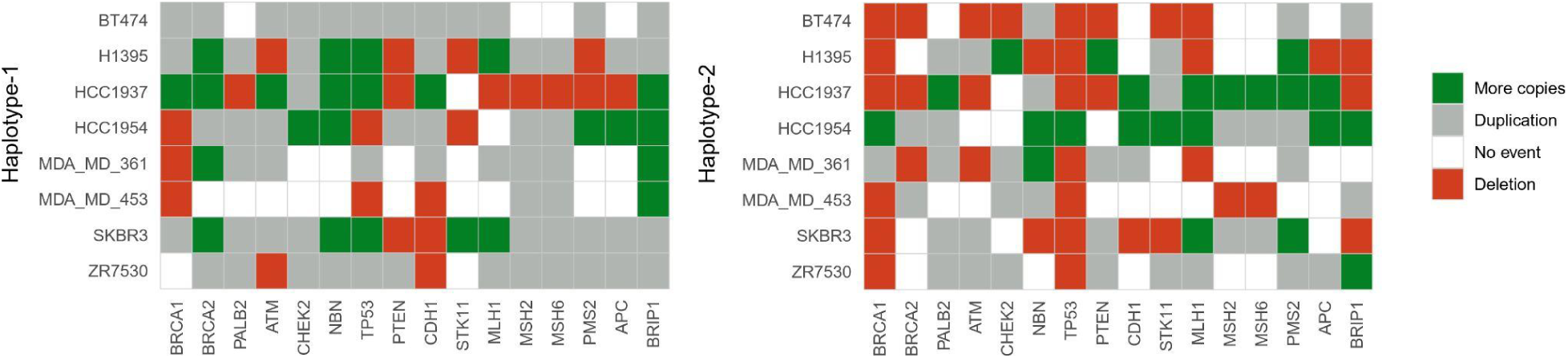
Haplotype-specific copy number profiling of breast cancer related genes in our breast cancer cell lines panel.

